# The Impact of Unemployment Benefits on the Mental Health of Underemployed Workers: Findings from the Understanding America Study

**DOI:** 10.1101/2025.10.16.25338149

**Authors:** Danny Maupin, Jungeun Olivia Lee, Autumn Kleinman, Woo Jung Lee, Haomiao Jin

**Affiliations:** School of Health Sciences, Faculty of Health and Medical Sciences, University of Surrey, Guildford, United Kingdom; Suzanne Dworak-Peck School of Social Work, University of Southern California, Los Angeles, CA; Institute for Addiction Science, University of Southern California, 1845 N Soto Street, Suite, 201C, Los Angeles, CA 90032; Keck School of Medicine, University of Southern California, 1975 Zonal Avenue, Los Angeles, CA 90033; UC Irvine Joe C. Wen School of Population & Public Health, University of California at Irvine, 856 Health Sciences Quad, Irvine, CA 92697, USA

**Keywords:** employment insecurity, depression, anxiety

## Abstract

**Background:** Underemployment, working fewer hours than desired, has a negative impact on mental health. Research suggests that unemployment benefits can support the mental health of unemployed individuals, but it is unknown if a similar benefit occurs for underemployed workers. This study aims to assess the relationship between unemployment benefits and mental health for underemployed workers.

**Methods:** Data was analysed from 18 waves (June 2020-March 2021) of the COVID-19 survey, a sub-study of the Understanding America Study. The dataset included responses from individuals (n=1,241) who self-identified as employed before COVID-19. A meta-regression created pooled effect sizes measuring the impact of unemployment benefits on the change in mental health of underemployed workers with two-, four-, six-, or eight-week effect lags.

**Results:** Receiving unemployment benefits is associated with a trend towards reduced depression at four weeks (−0.13, 95%CI −0.25; 0.00) and reduced depression six weeks later (−0.14, 95%CI −0.23; −0.05). A significant reduction in anxiety scores was seen six weeks after receiving unemployment benefits (−0.18; 95%CI −0.34; −0.01). Results from gender were mixed with female workers noting a positive effect from unemployment benefits, and male workers showing no or even negative effects, particularly on anxiety.

**Conclusion:** These findings underscore the impact of unemployment benefits on underemployed workers’ mental health. Unemployment insurance, not a typically utilized policy strategy to improve mental health, may mitigate mental health repercussions of underemployment. Expanding benefits to include underemployed workers should be considered particularly during societal crises such as COVID-19.

**What is already known on this topic:** It is known that underemployment and employment insecurity can have negative impacts on mental and physical health, however it is not known whether providing benefits to underemployed individuals.

**What this study adds:** We now know that adding benefits to underemployed works can improve depression and anxiety, particularly in female workers.

**How this study might affect research, practice or policy:** This provides avenues for future research to address the impacts of benefits for underemployed workers, particularly for assessing causality. Further, it provides initial evidence that can be used to expand policy by including underemployed workers within unemployment benefits.

## INTRODUCTION

Mental health disorders are a growing public concern across multiple countries. In the United States, compared to 1993-1999, twice as many young adults and 50% more of adults in 2018-2020 over the age of 25 meet the criteria for moderate to severe depression (1). While differences were found between gender (e.g., males experiencing a larger increase compared to females) and race (larger increases in White Americans than Black Americans), increases in mental distress were seen across all demographic groups (1). Mental health problems have been linked to negative physical health outcomes - approximately 16% of disability-adjusted life years could be attributed to mental disorders in 2019 (2) – and economic costs – estimated to be 282 million USD in the US alone (3). Thus, identifying factors contributing to mental health and developing effective public health policies are crucial to ensuring healthier lives of the general population while also reducing economic and government costs.

Employment insecurity is a critical factor shaping mental health, with a robust field of evidence supporting this link and documenting its scarring effect. For example, a study of 201 countries across 50 years of data found a significant link between unemployment and mental health conditions including depression and anxiety (4). Similarly, another meta-analysis found that job insecurity was associated with depression, anxiety, and emotional exhaustion (5). Our own research further highlights that not only do downward transitions in employment security, such as from full time employment to unemployment, result in increased symptoms of depression and anxiety, but regaining job security also did not immediately lead to improvements in these symptoms (6).

Although providing important insights into the link between employment insecurity and mental health, existing studies are limited in two critical ways. First, there is a conventional assumption that having any job is better than none. As such, underemployment—individuals who are employed but would like to work more hours than they currently do (7)–has often been erroneously lumped with secure full-time employment erroneously. Employment is not a dichotomous outcome (e.g., working vs not working); rather it exists on a continuum ranging from adequate to inadequate employment (8). While estimates vary the number of underemployed workers in the United States have been reported to be 4.6 million in September 2018 – compared to ∼6 million unemployed – and a peak value of 9 million in September 2011 – compared to 15.4 million unemployed – suggesting a tendency for underemployment to rise during economic downturns (9,10). Underemployed workers have been often overlooked in existing studies, despite making a significant proportion of the workforce. In a few studies that focused on underemployment, underemployment showed a similar impact on depression as unemployment (8). Part time work has been found to have negative effects on stress and health behaviours including healthy eating and cigarette smoking (11).

Second, it is unknown whether income supplementation at the event of underemployment can mitigate its adverse impacts on mental health. For unemployed individuals, some of income loss triggered by job loss can be recovered via unemployment insurance benefits. Confirming such conceptual speculation, previous research has found that unemployment benefits mitigate the impacts of unemployment on depression (12). A systematic review by Renahy et al. (13) identified unemployment benefits have been found to have positive effect on a range of outcomes including trend towards protective effects on mental health, protective effects on poverty, and subjective well-being. An analogous inquiry in the context of *under*employment has been absent in U.S., because underemployed individuals are typically not eligible for unemployment insurance. During Covid, the unemployment benefits were expanded in the U.S. to underemployed workers, a segment of workers who are not typically included in the employment insurance program (14). As such, this period presents a unique opportunity to assess whether employment insurance mitigates mental health repercussions of underemployment.

Finally, prior studies have shown that the damaging impacts of employment insecurity may vary by demographics, particularly by gender. In one study, women and younger workers have been shown to more likely to suffer negative impacts of underemployment (15), while males and those over the age of 45 were reported to have heightened odds of experiencing depression or anxiety in another study (16). Adding to the confusion, a study by Rosenthal et al. (11) found no significant differences across age, race, education, or gender have been reported. Gender differences across unemployment and impacts on mental health have been documented in the literature before, with various reasonings provided. For *un*employed workers, it was found that marriage and social class provided different effects on mental across gender. Marriage decreased mental health in unemployed men, but acted as a buffer for unemployed women, while men who lost a manual job saw larger decreases in mental health compared to the opposite effects in women (17).

To our best knowledge, no studies have evaluated gender differences in the protective effects of unemployment insurance in underemployed workers. Yet such differences are likely, given that historically disadvantaged social groups —such as women (18), racial ethnic minority groups (19), and low socio-economic status (20,21)—often have limited resources (19). For these groups, the income support provided through unemployment insurance may play a particularly important protective role and thus understanding the role of unemployment insurance in shaping mental health among underemployed workers is critical for informing effective policy programming.

### Current Study

The current study aims to explicitly focus on *under*employed individuals, a highly prevalent yet overlooked group affected by employment insecurity. Specifically, the study will assess the impact of receiving unemployment benefits on the mental health of underemployed individuals during the COVID-19 pandemic when eligibility to unemployment insurance had been expanded to include underemployed individuals. Additionally, this study will assess how these benefits will impact individuals over a two-to-eight-week period, an important consideration noting a relationship between timing of benefit provision and mental health outcomes (6), and across gender.

## METHODS

Data for this study was obtained from the Understanding America Study (UAS). The UAS is an American population-based internet panel of adults aged 18 years and older, recruited using probability-based approaches drawing participants based on their address (22).

This recruitment is conducted in batches, with 34 batches of recruitments completed between 2014 to 2024. Further information, such as the proportion of individuals who received invitations and retention rates by batches can found on the UAS website (23).

For the purposes of this study, the biweekly COVID-19 tracking surveys were used. These surveys were conducted from April 2020 to March 2021, totaling 24 waves over the course of the year and covering the peak unemployment rate and subsequent decline over this period. Over 7000 of approximately 9500 UAS members agreed to participate (24). The response rate was over 90% in earlier waves and remained above 60% throughout the majority of the survey period. For this study, only participants who experienced underemployment were included. Underemployment is defined as individuals who experienced involuntary reduction in the number of working hours, typically referred to as hour-related underemployment (25). This was inclusive of 1) people who reported a reduction in their working hours compared with prior waves or 2) who reported working fewer than 35 hours or their pre-COVID-19 work hours. This definition is consistent with the International Labor Organization’s (ILO) (26) definition. Further, only data from Wave 7 (survey open June 10 - July 8, 2020) to Wave 24 (survey open February 2, 2021 - March 3, 2021) are utilised as employment benefits were not broadly offered to underemployed individuals until Wave 7 in U.S. This resulted in a final sample of n=1241 (See Figure 1 for the derivation of the analytic sample).

**Figure 1.**
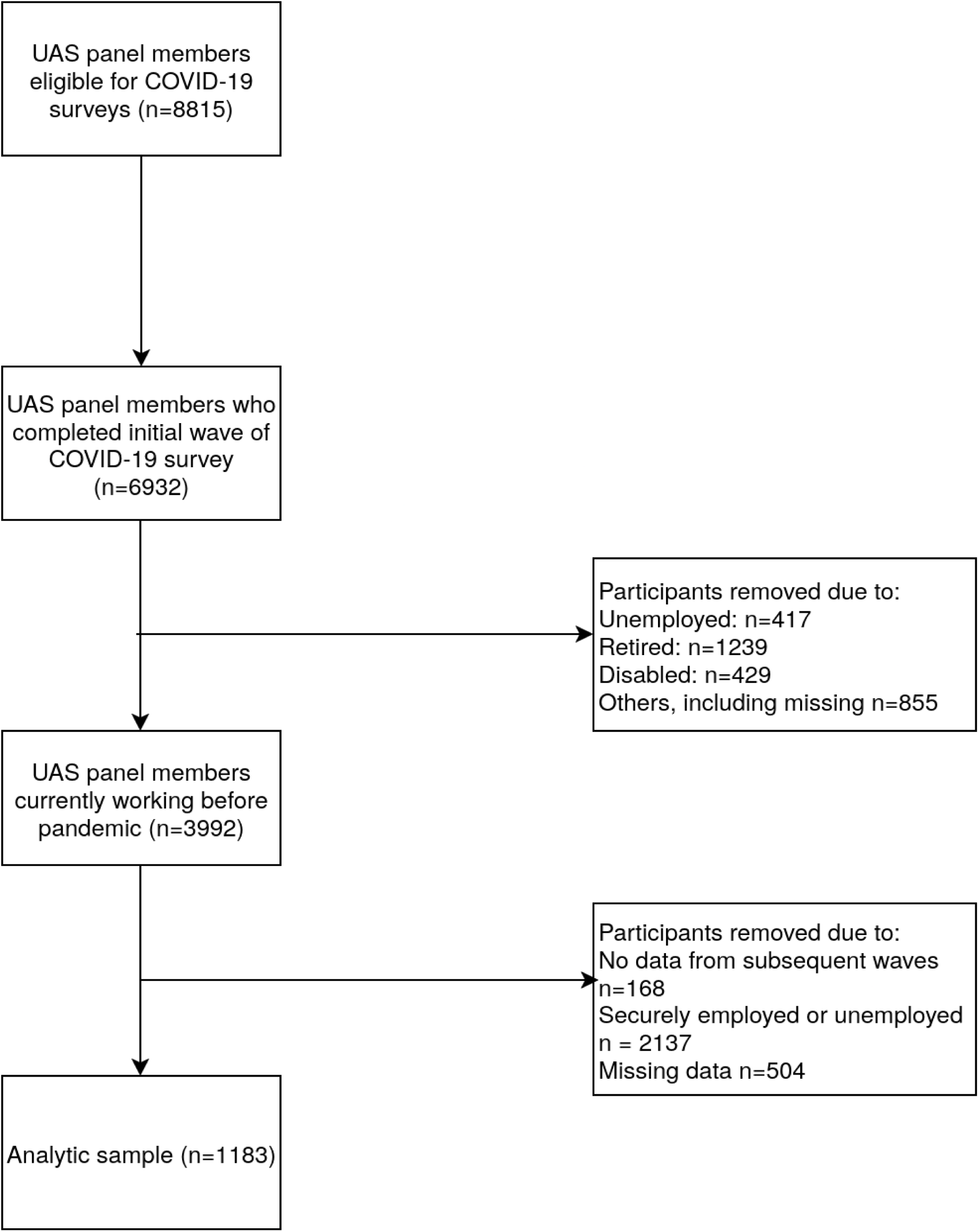
Selection of analytic sample, UAS – Understanding America Study

### Measurements

#### Mental Health

Two aspects of mental health were assessed, depression and anxiety. Depressive and anxiety symptoms were measured with the Patient Health Questionnaire-2 (PHQ-2) (27), and Generalized Anxiety Disorder scale (GAD-2) (28), respectively. Both scales have shown widely established reliability and validity (29).

We assessed the change in PHQ-2 and GAD-2 scores subsequent to underemployment across 4 different time points: 2 weeks (next wave) later, 4 weeks (two waves) later, 6 weeks (three waves) later, and 8 weeks (four waves) later. These time points were chosen to assess the effects of any time lag between the provision of unemployment benefits and the impact on mental health.

#### Sociodemographic Variables

We further assessed the relationship between unemployment benefits on underemployed individuals across gender. Differences across race and education were not assessed due to smaller sample sizes across groups resulting in likely low power to result in meaningful analysis.

### Statistical Analysis

#### Overall

A range of statistical techniques has been implemented using R. First, missing data was handled using multivariate imputation by chained equations via the mice package (30) (see the online supplemental material 1 for more details about proportion of missingness). Next, statistical matching was conducted to control for multiple confounders between individuals who received unemployment benefits compared to those who did not across each wave using the MatchIt package (31). Confounders included demographic characteristics (e.g., age, race, education), household information (e.g., income, number of family members), substance use (e.g., binge drinking), mental health history, and perceived risks during the Covid-19 pandemic (e.g., perceived risk of dying, infection) (see the online supplemental material for a full list of variables used). Third, a regression analysis was deployed to predict the outcome (i.e., change in mental health scores) in relation to receiving unemployment insurance among underemployed workers, while controlling for age, race, gender, education, marital status, household income, number of household members, cannabis use, and substance use. This regression equation was completed independently at each wave of surveys during the study period (June 10, 200 – March 3, 2021). Meta-regression was then used to create a pooled effect size across all waves during the survey periods. Random effects models, which assumes the true effect varies across comparisons rather than the single true effect assumed by fixed effects models, were chosen due to the high heterogeneity across all comparisons (*I^2^* > 60% across all comparisons). Meta-regression was chosen for statistical analysis as the number of underemployed workers, and those receiving unemployment benefits, are quite small compared to the rest of the sample at each wave and thus difficult to achieve a stable estimate across all statuses (6), particularly with more robust statistical methods such as marginal structural models (32).

#### Gender specific analysis

To assess the relationship between unemployment benefits and mental health in various demographics group, we utilised a stratified analysis whereby we implemented the same imputation, matching, and meta-regression across each gender. However, due to smaller sample sizes, fewer covariates had to be used in these stratified analyses, compared to the initial matching for the full analysis sample. The covariates were limited to the following: age, marital status, education, race, hours worked, household income, and number of household members. Gender was not included as a matching covariate as all participants would have the same value.

## RESULTS

A breakdown of the sample characteristics and demographics is shown in Table 1.

**Table 1.** Demographic characteristics of analytical sample.

| Demographic characteristic | Mean (± SD) or N (%) |
| --- | --- |
| Age (years) | 45.4 ± 13.3 |
| Gender |  |
| Male | 461 (37.1%) |
| Female | 781 (62.9%) |
| Education |  |
| High school or less | 241 (19.4%) |
| Some college | 468 (37.7%) |
| Bachelor's degree | 298 (24.0%) |
| Graduate degree | 235 (18.9%) |
| Race |  |
| White | 934 (75.2%) |
| Black | 116 (9.3%) |
| Other | 192 (15.4%) |

### Overall data set

#### Depression

Figure 2 shows the meta-regression forest plot across the four time periods in the overall dataset with depression as an outcome. In this instance, negative estimates would describe a decrease in depression and anxiety when provided unemployment benefits.

**Figure 2:**
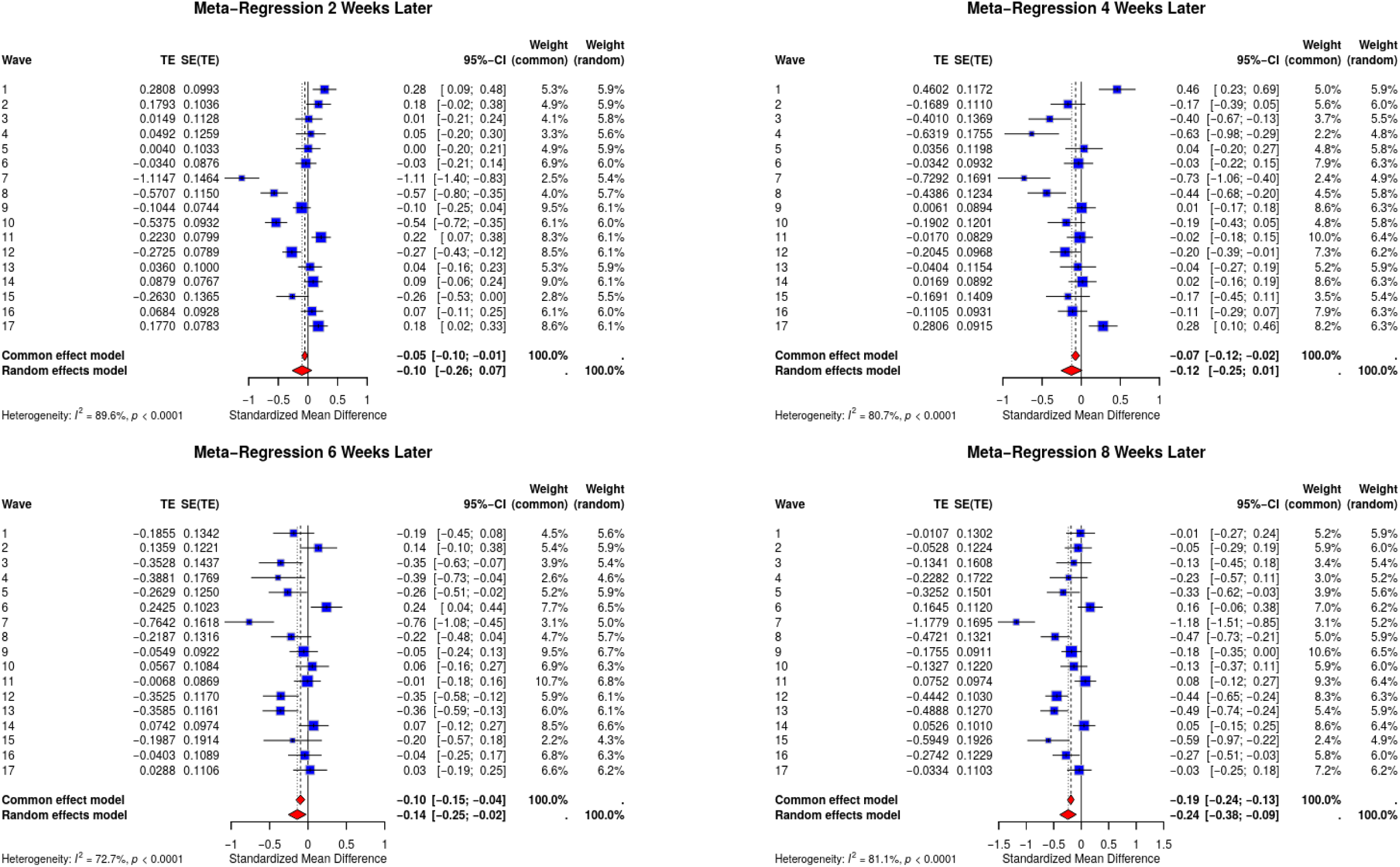
Meta-regression forest plots depicting outcome of unemployment benefits on depression as measured by PHQ-2 scores Note: In clockwise order outcomes are as follows - a) two weeks later, b) four weeks later, c) eight weeks later, d) six weeks later

Receiving unemployment benefits was associated with no significant change in depression scores two weeks later in the random effects model (REM) (−0.10; 95% CI - 0.26; 0.07). However, the receipt of unemployment benefits is associated with a trend towards a reduction in depression at four weeks (−0.12, 95% CI −0.25; 0.01) and reduced depression six weeks (−0.14, 95%CI −0.25; −0.02), and eight weeks (−0.24, 95% CI −0.48; −0.09) post receipt. This suggests that for the overall sample of underemployed workers, providing unemployment benefits significantly reduced reported depression with a time delay of four weeks.

#### Anxiety

Figure 3 shows the meta-regression forest plot across the four assessment points in the overall sample with anxiety as the outcome. Receiving unemployment benefits have no significant impact on anxiety scores two weeks (−0.03; 95% CI −0.14; 0.07) and four weeks later (−0.11; 95% CI −0.27; 0.03). However, six weeks after receiving unemployment benefits, there is a significant reduction in anxiety scores (−0.18; 95% CI −0.34; −0.01), which increases by eight weeks post-receipt (−0.20; 95% CI −0.34; −0.05). Similar to depression, providing unemployment benefits to underemployed workers significantly reduced reported anxiety. However, there was a longer time delay of six weeks for the protective impacts.

**Figure 3:**
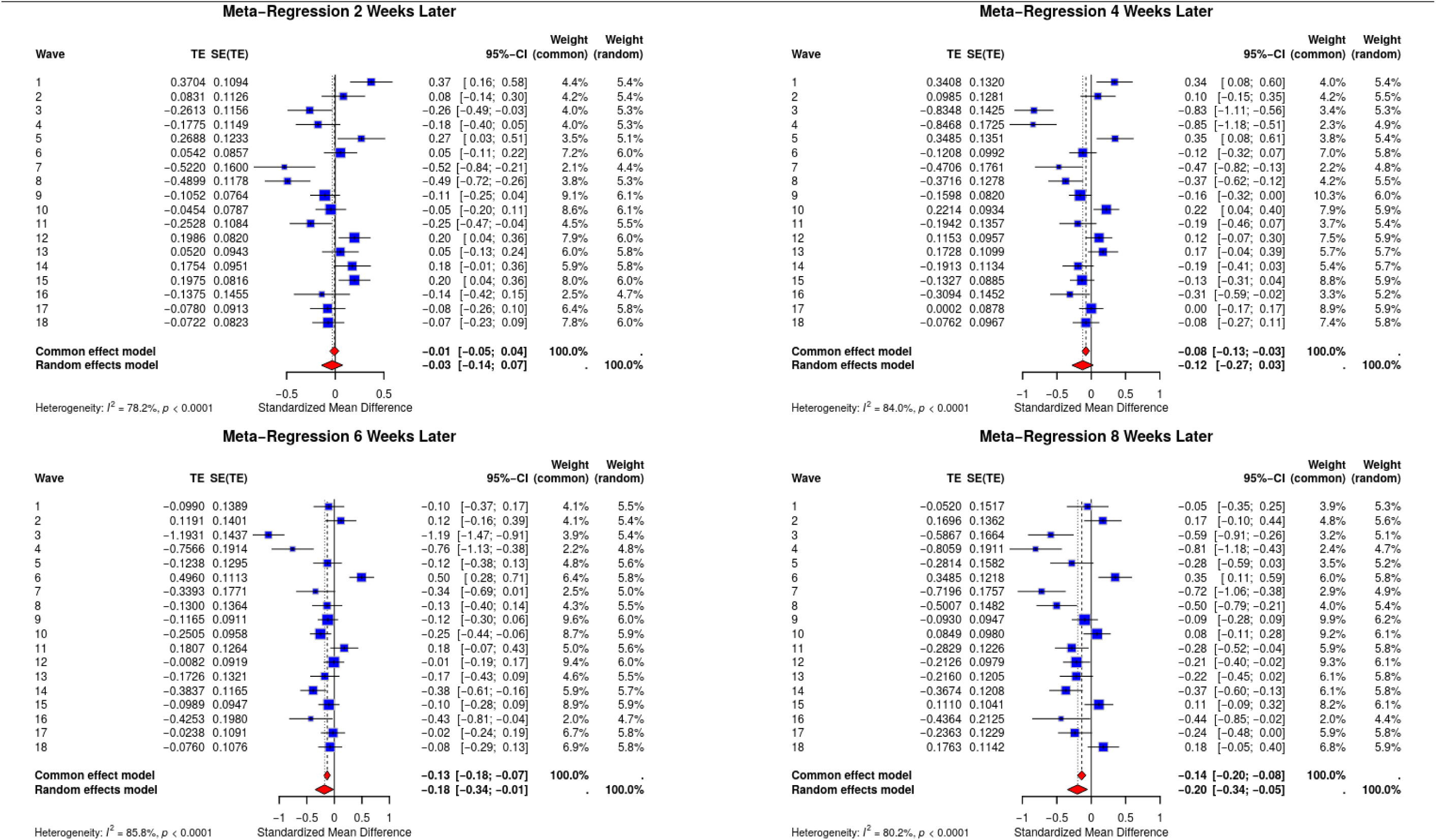
Meta-regression forest plots depicting outcome of unemployment benefits on depression as measured by GAD-2 scores Key: In clockwise order outcomes are as follows - a) two weeks later, b) four weeks later, c) eight weeks later, d) six weeks later

### Gender

#### Depression

Next, we evaluated the impact of unemployment benefits on depression for underemployed workers across genders. The pattern in the impact of benefits on depression scores in female participants is like the one observed in the full sample but with larger effect sizes. There is a trend towards decreased depression scores two weeks later after receiving unemployment benefits (−0.15, 95% CI −0.34; 0.05). Significant reductions in depression have been also observed at four (−0.26; 95% CI - 0.46; −0.05), six (−0.33; 95% CI −0.50; −0.15) and eight weeks (−0.35; 95% CI −0.53; - 0.17) post receipt of benefits. Males in this sample experienced a different pattern with no effect occurring in two weeks later (0.00; 95% CI –0.13; 0.14) and then a trend towards *increased* depression scores four (0.10; 95% CI –0.03; 0.22) and eight weeks later (0.11; 95% CI –0.05; 0.27), and significant increases six (0.14, 95% CI 0.05; 0.24) weeks post receipt, suggesting a possible difference in response to receiving unemployment benefits across genders. Meta-regression forest plots from these stratified analyses can be found in Supplementary File 2.

#### Anxiety

Among female participants, receiving unemployment benefits did not translate into any change in anxiety scores two weeks (−0.06; 95% CI −0.24; 0.12) or four weeks (−0.23; 95% CI −0.48; 0.02) post-receipt, despite a decreasing trend. However, receiving unemployment was associated with a significant decrease in anxiety at six (−0.28; 95% CI −0.51; −0.05) and eight weeks (−0.31; 95% CI −0.50; −0.11) post-receipt. For male participants unemployment benefits resulted in significant increases in anxiety across all time points: two (0.13, 95% CI 0.05; 0.22), four (0.19, 95% CI 0.06; 0.33), six (0.22, 95% CI 0.05; 0.39), and eight (0.22, 95% CI 0.07; 0.38) weeks post receipt.

## DISCUSSION

The aim of this research was to assess the impact of unemployment benefits on the mental health of individuals who are underemployed, and how these benefits vary across varying sociodemographic populations during the COVID-19 pandemic. Overall, these results suggest that providing unemployment benefits to underemployed workers has a significant impact on their depression and anxiety. These impacts do not appear to emerge immediately after receiving unemployment insurance. Rather, protective effects surface six weeks after the provision of unemployment benefits for both depression and anxiety.

Unemployment has been shown to have a negative impact on an individual’s mental health, with additional research suggesting these impacts are similar between un- and under-employed individuals (33). Although unemployment benefits have been shown to mitigate these negative effects in unemployed individuals (12), little is known whether receiving income support via unemployment insurance can generate protective effects among underemployed individuals. The findings in this paper suggest that expanding unemployment insurance eligibility to underemployed individuals may reduce mental health problems benefiting an individual’s well-being and potentially leading to cost savings at a society level. In the US, the cost of mental illness is estimated to be 217 billion USD when accounting for impaired function in the work place, healthcare expenditures, and suicide-related costs, however estimates can reach up to 282 billion USD when accounting for other factors such as accounting for savings and potential implications on job choice (3). Research on the impact of unemployment benefits to the mental health of part-time workers is scarce. Broadly, two studies by Kyyrä (34,35) found that the addition of underemployment benefits reduced the time for individuals to return to full-time work. An earlier return to full-time work, in addition to supplemental income, is one potential mechanism for the improved mental health outcomes as full time workers tend to have better health outcomes (11).

Consistent with prior studies, our findings suggest that unemployment insurance, not typically considered as a population-level mental health intervention, may have the potential to mitigate psychological repercussions of underemployment. As underemployed workers are not typically eligible for unemployment insurance, our findings suggest further research and exploration on expanding the unemployment insurance.

These results are suggestive of a time-sensitive component, with the improvement of unemployment benefits strongest at six weeks later for both depression and anxiety. This echoes previous research which found detrimental effects of unemployment on mental health did not immediately subside with the addition of benefits (6). Coupled with the prior study (6), our findings suggest that the restorative effect of unemployment insurance may not be immediate, but instead require a latency period before their protective effects on mental health emerge to surface. As there appears to be a time lag for the impacts of unemployment insurance to be felt, it is imperative that benefits are provided in a timely manner to begin to recover from possible scarring effects on their mental health as soon as possible (36). Improving the efficiency of the claiming process will assist in minimizing depression and anxiety in workers and helping them to remain an effective part of the work force.

These results were inconsistent across demographic groups. Female workers followed a similar pattern with trends towards decreases that became significant six and eight weeks later for both depression and anxiety. On the other hand, male workers saw significant *increases* in anxiety across all time points and significant increase in depression at six weeks post benefit receipt. Interestingly and in possible contrast to our work, Kyyrä (34) found that subsidised part-time work significantly reduced the time to return to full time work for male but not for female workers. Alternatively, it could also be results of lower sample sizes in the demographics, with female workers (n=781) are almost double that of male workers (n=481), a not unusual finding as previous research has shown that female workers may be more likely to be underemployed (37). Sample sizes were more balanced in previous research finding males suffered greater mental health effects from unemployment (57.8% female; 42.2% male) (16) and research showing females suffered greater mental health effects (51.2% female; 48.8% male) (15). Further contributing to the complex interactions of gender, mental health, and employment security are the effects of marriage and social status. As mentioned in the introduction, these variables show opposite effects across gender and are potentially influenced by status of sole-providers, traditional provider views for genders (e.g., nurturant or breadwinner roles) (17). In the current study, it is possible that male anxiety rose while on benefits due a to perceived inability to fulfill their role. Future studies can seek to understand this in greater detail by adding questions on participants and their perceived roles to evaluate the impacts these have on depression and anxiety in underemployed workers.

This research is not without limitations. First, this analysis is unable to establish a causal relationship between the receipt of unemployment benefits and improvements in mental health due to the observational nature of the data, likewise though we utilised mental health history in statistical matching, it is not our intention to claim that such approach completely rules out any reverse causation possibility. Similarly, although we have employed statistical strategies (i.e., propensity matching) to mitigate this concern, any causality claim should be cautioned. Of note, we also considered another statistical modeling strategy known to further mitigate this concern including marginal structural models (32,38). However, data sparsity across the different data periods led to estimation challenges. Future research on a sample with a keen focus on underemployment may be a fruitful future direction allowing for statistical methods to assess causality. Secondly, our underemployment measure did not account for income- or skills-based underemployment (39), rather focusing on hours-based underemployment potentially underestimating the effects of underemployment on mental health and the subsequent effect of unemployment benefits. Additionally, other potential biases may exist from the use of self-reported data on work hours (40), a bias towards individuals with higher digital literacy given the dataset is an internet-based panel.

## CONCLUSION

This study utilised a national, multi-wave survey to assess the impact of unemployment benefits on the mental health of underemployed workers. Our study represents, to the authors’ knowledge, the first to investigate this relationship, revealing that unemployment insurance benefits have resulted in a significant reduction in depression and anxiety. This relationship was not consistent across all sociodemographic groups with female workers seeing positive impacts from underemployment benefits, while male workers showed no significant or even negative impacts. These results highlight the potential impact of unemployment benefits for underemployed workers, providing possible policy options that continue with the expanded unemployment benefits to include underemployed workers to provide mental health benefits, particularly for female underemployed workers.

## Acknowledgements

The UAS was supported by the Bill & Melinda Gates Foundation and a grant from the National Institute on Aging (5U01AG054580).

## Data availability statement

Data are available in a public, open-access repository. The data used for this study are publicly available on the UAS website (https://uasdata.usc.edu/index.php).

## Ethics statements

### Patient consent for publication

Not applicable.

### Ethics approval

This study used anonymised public data and did not require ethics review. The UAS was approved by the University of Southern California Institutional Review Board. Participant consent was obtained during original data collection.

### Contributors

DM, HJ, JOL and WJL contributed to the conceptualisation and design of the study. DM and HJ handled data curation and analysis. DM and AK drafted the paper, and all authors contributed to its critical review and editing. DM is responsible for the overall content as guarantor.

### Disclaimer

The funding agencies played no role in any aspects of the present study, including the design, data collection, data analysis, and interpretation of the study and the decision to submit the study for publication.

