## Supplemental File 1 for "The Impact of Unemployment Benefits on the Mental Health of Underemployed Workers: Findings from the Understanding America Study"

Supplemental Table: Variables used in matching and respective percent missingess in included variables

| Variable | Percent missingness |
| --- | --- |
| PHQ2 | 0% |
| GAD2 | 0% |
| Receive unemployment insurance | 0% |
| Gender | 0% |
| Age | 0% |
| Marital status | 0% |
| Education | 0% |
| Race | 0% |
| Hours work | 4.9% |
| Household income | < 1% |
| Number of household members | 3.1% |
| Cannabis use | 0% |
| Drug use | < 1% |
| cr056a | < 1% |
| cr056b | < 1% |
| cr056c | < 1% |
| cr056d | < 1% |
| cr056e | < 1% |
| cr056f | < 1% |
| cr056g | < 1% |
| cr056h | 1.4% |
| cr056i | 0% |
| cr054s1 | 0% |
| cr054s2 | 0% |
| cr054s3 | 0% |
| cr054s | 0% |
| cr054s5 | 0% |
| cr054s6 | 0% |
| cr054s7 | 0% |
| cr054s8 | 0% |
| cr054s9 | 0% |
| cr054s10 | 0% |
| cr054s11 | 0% |
| lr011 | 21.5% |
| Perceived risk of dying due to Covid | < 1% |
| Perceived risk of Covid infection | < 1% |
| Perceived risk of job loss | 19.8% |
| Perceived risk money | < 1% |
