## Supplementary figures and images for "The Impact of Unemployment Benefits on the Mental Health of Underemployed Workers: Findings from the Understanding America Study"

### Supplemental File 2

#### Meta-regression forest plots for gender

##### Depression

###### Female

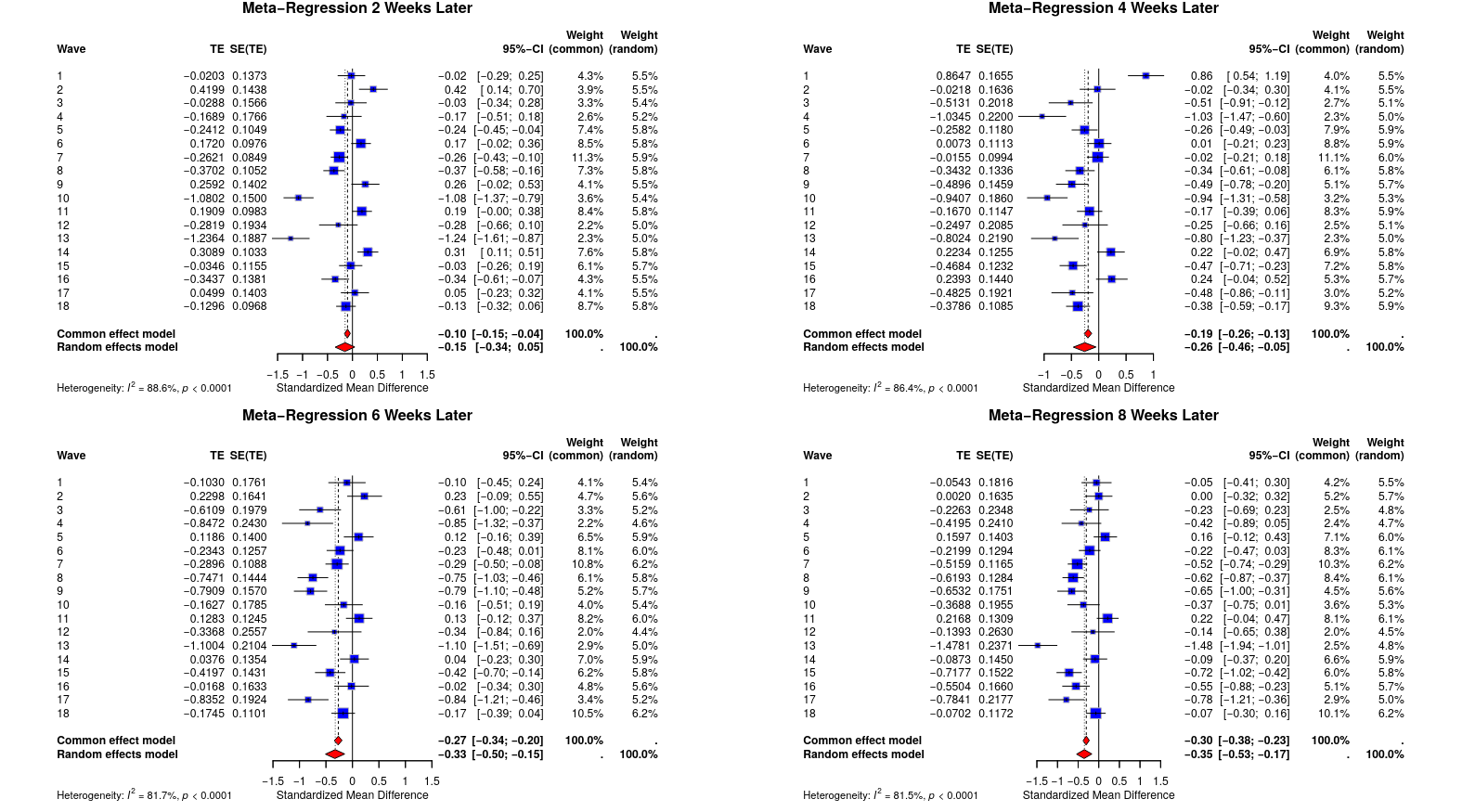

Male

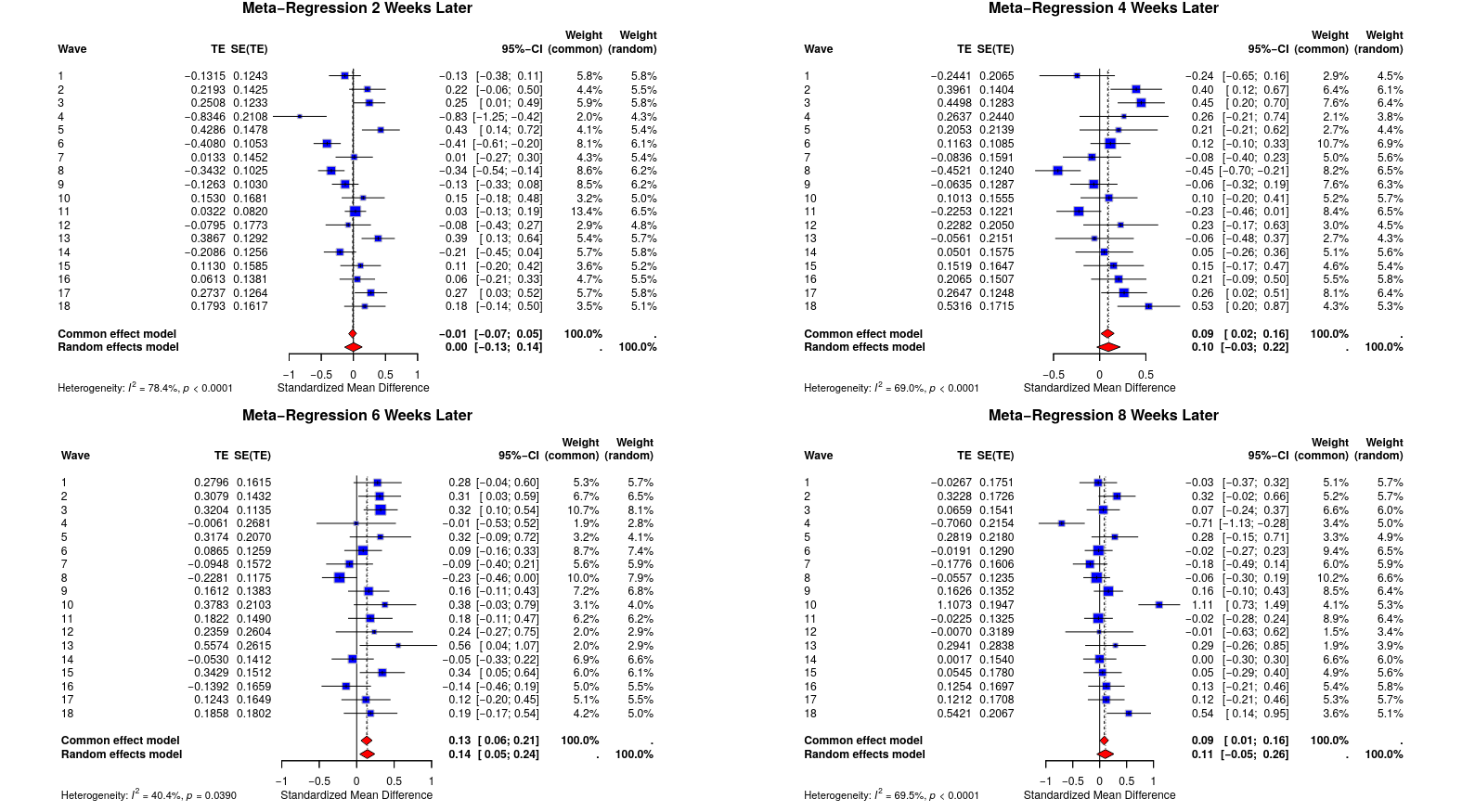

##### Anxiety

###### Female

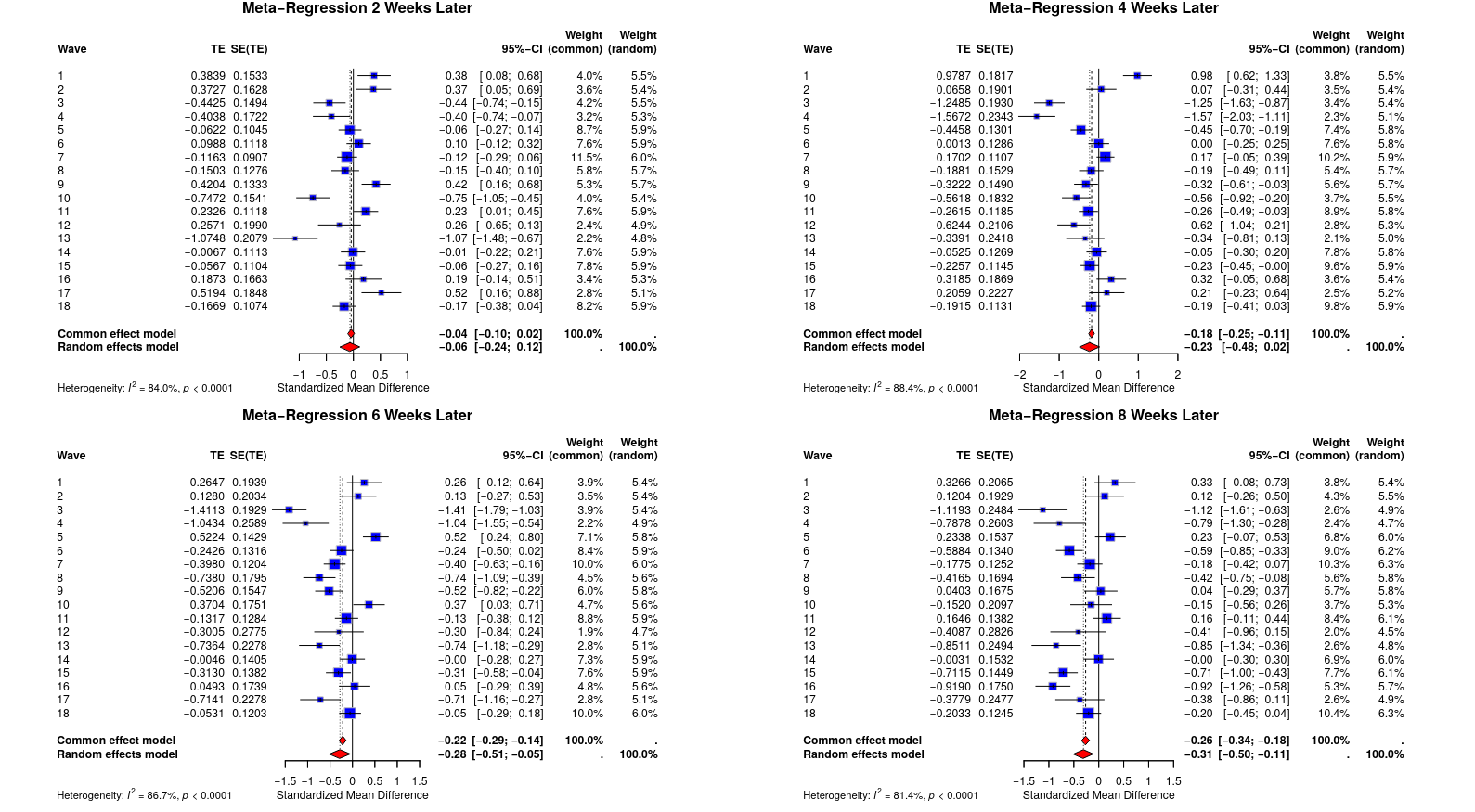

###### Male

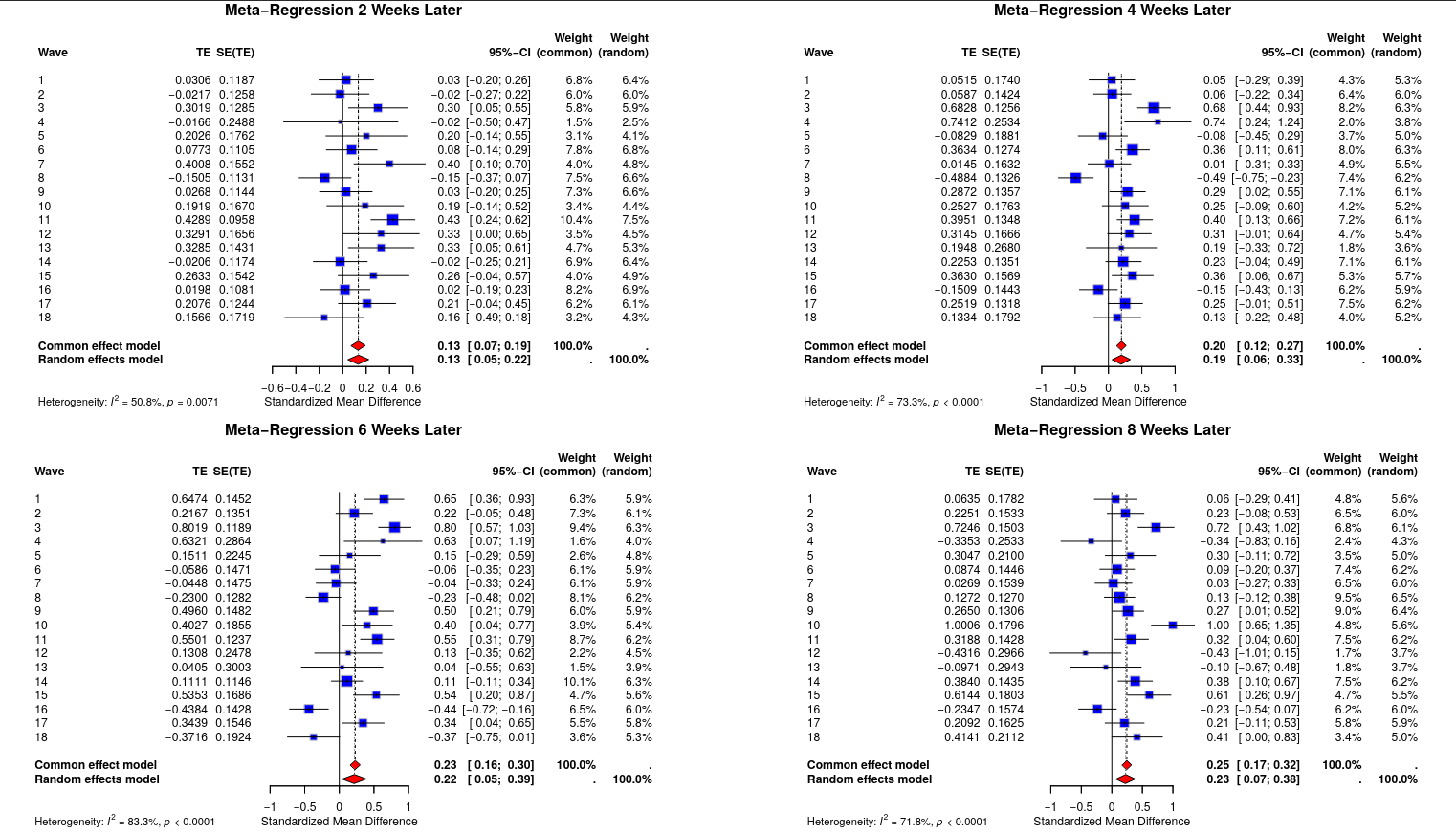
